# Semaglutide and non-arteritic anterior ischaemic optic neuropathy: a 121-fold reporting signal and a bounded genetic null

**DOI:** 10.64898/2026.09.06.26362369

**Authors:** Junhong Jiang, Di Hu, Xinsheng Li, Qipeng Ling, Qi Zhang, Lili Dong, Zenan Lin, the μ-Biomedical Data Investigation Group (Mu-BioDig)

## Abstract

**Background:** A cohort study on 3 July 2024 linked semaglutide to non-arteritic anterior ischaemic optic neuropathy (NAION), and in June 2025 the European Medicines Agency (EMA) accepted NAION as a very rare side effect, about one additional case per 10,000 person-years.

**Objective:** To measure how much of the signal the reporting system generates itself, test whether three reporting systems are independent, and let a genetic instrument adjudicate rather than corroborate.

**Methods:** FAERS (12,305,041 reports, 2018Q1 to 2026Q2), JADER (1,054,239 reports), and VigiAccess counts were analysed with reporting odds ratios, covariate-adjusted regression, and pre-specified probes. Six cis-eQTL variants for GLP1R expression instrumented anterior ischaemic optic neuropathy (AION, 244 cases) and five AION-spectrum traits, with colocalisation.

**Results:** Semaglutide carried 879 NAION reports and a reporting odds ratio of 120.8 (95% CI 110.6 to 132.0), about sixty times the relative risk the EMA accepted, and the odds that a report named NAION rose 48.6-fold afterwards. VigiBase held 871 such reports where FAERS held 879, and the two systems give opposite accounts of the GLP-1 class. Of fifteen observations separating toxicity from reporting dynamics, one favoured reporting dynamics, two favoured toxicity weakly, two bounded an expression-mediated mechanism, and ten decided nothing. GLP1R expression was unassociated with AION (0.56, 0.21 to 1.49), a null powered only above fourfold, while glaucoma was bounded (1.05, 0.87 to 1.25) and no relevant ocular trait colocalised.

**Conclusions:** Semaglutide’s NAION signal is amplified by the reporting system, and the databases that appear to corroborate it hold the same records, so the causal question stays open in both directions.

## 1. Introduction

Non-arteritic anterior ischaemic optic neuropathy (NAION), the most common acute optic neuropathy in adults over fifty, causes sudden, painless, irreversible loss of vision.

On 3 July 2024 a retrospective cohort study reported a four-to sevenfold higher rate of NAION among patients prescribed semaglutide [1], and Danish and Norwegian register studies followed with an estimate near twofold [2]. In June 2025 the EMA’s Pharmacovigilance Risk Assessment Committee (PRAC) concluded that NAION is a very rare side effect of semaglutide medicines, roughly one additional case per 10,000 person-years of treatment [3], and the World Health Organization issued a global alert the same month [4].

Stimulated reporting, notoriety bias, and case dredging produce large disproportionality estimates with no pharmacological cause [5, 6], and a genuine toxicity arrives through the same channels and looks similar.

Two literatures have read this pair in opposite directions. Meta-analyses pool the observational estimates and one puts the word induced in its title [7–9], while a critique argues that no underlying design supports a causal claim [10]. Multi-database disproportionality studies treat the agreement between FAERS and VigiBase as confirmation [11–13], without stating the gap between reporting and epidemiology and without testing whether the systems they triangulate are independent. Pairing a reporting database with a genetic proxy for the same target is established practice [14–16], and there the genetic arm corroborates rather than risks the verdict.

We measure the gap instead, treating a rare toxicity amplified by awareness and reporting dynamics without toxicity as competing explanations and asking which of them these data can decide.

## 2. Methods

### 2.1 Design and data sources

The study has five components: disproportionality and reporting-dynamic analysis in FAERS, a second analysis in JADER, aggregate counts from WHO VigiBase through VigiAccess, descriptive use of the European regulatory assessment, and drug-target Mendelian randomisation (MR) with colocalisation at the GLP1R locus (Figure 1).

**Figure 1.**
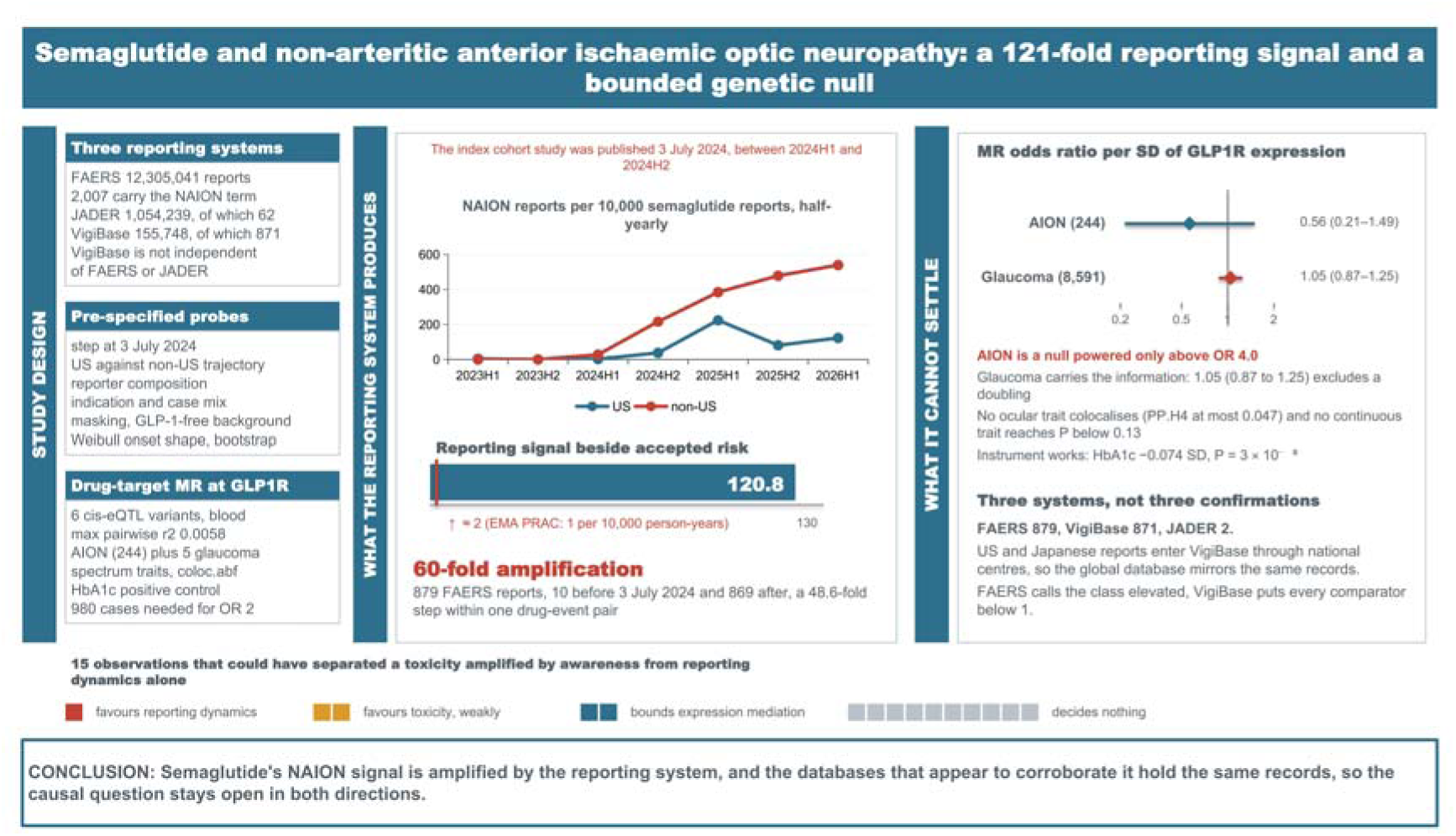
The workflow.

FAERS covers 2018Q1 to 2026Q2 from quarterly ASCII extracts, deduplicated to the most recent version of each case, giving 12,305,041 unique reports. JADER covers 2004 to 2026 and gives 1,054,239 reports, and VigiAccess counts were retrieved on 22 October 2026 [17]. The reference date for the alert is 3 July 2024 [1]. FAERS analyses use the whole database as the primary background and a study-drug universe of GLP-1 receptor agonists, SGLT2 inhibitors, DPP-4 inhibitors, metformin, and atorvastatin as a conservative second background. National centres including the FDA and the PMDA contribute to VigiBase [18], so the VigiBase arm is not independent of FAERS or JADER.

### 2.2 Outcome and exposure

NAION has no dedicated MedDRA preferred term. The relevant lowest-level terms map onto one preferred term, “Optic ischaemic neuropathy” (PT code 10030924), an EMA designated medical event, and that term is the primary outcome. In JADER the term is 虚血性視神経症, and VigiAccess displays “Ischaemic optic neuropathy”. Drug groups were assigned by regular-expression matching of generic and brand names in the primary-suspect role in FAERS and the suspect-drug role (被疑薬) in JADER. A four-term sensitivity set adds the anterior, generic, and non-arteritic forms.

### 2.3 Disproportionality

Reporting odds ratios (ROR) and information components were computed with Haldane 0.5 correction and Norén shrinkage within each background stratum [6, 19], with Benjamini– Hochberg correction inside each background by outcome family. A signal was flagged when the lower 95% bound exceeded 1 and the drug had at least three cases. Adjusted RORs come from logistic regression of NAION status on drug group with age group, sex, report year, US region, reporter type, and indication class as covariates against metformin as reference. A model without report year is also reported, since year adjustment partly absorbs the post-2024 surge. VigiAccess exposes only aggregate counts, so RORs there are graded for direction only.

### 2.4 Reporting-dynamic probes

Each probe was pre-specified and is interpretable under both hypotheses. We examined regional structure, time to onset, reporter composition, case mix and indication, masking against a background with all GLP-1 reports removed, the post-alert non-US trajectory by reporter type, and a bounding analysis of Americas reporting density. Full probe definitions and the two Weibull fitting routes are in Supplementary Methods.

### 2.5 Drug-target MR and colocalisation

The exposure is cis-genetic regulation of GLP1R expression in whole blood, from the eQTLGen dataset eqtl-a-ENSG00000112164 (hg19) [20]. Because all 205 candidate variants in the retrieval window fall inside one 340 kb region, LD pruning was done locally with PLINK 2.0 against the LDSC 1000 Genomes EUR Phase 3 chromosome 6 fileset at r² below 0.01, and the set was rebuilt on a second machine with a separate binary and reference-panel copy, giving identical instruments and an identical LD matrix.

Outcomes came from OpenGWAS [21]: AION (finn-b-DM_AION, 244 cases and 218,051 controls of European ancestry) [22] as the closest available proxy for NAION, then intraocular pressure, vertical cup-disc ratio, retinal nerve fibre layer thickness, ganglion cell inner plexiform layer thickness, and glaucoma, with HbA1c as positive control and ease of skin tanning as negative control. Inverse-variance weighting is primary, with weighted median and MR-Egger as sensitivity analyses. coloc.abf [23] was run over chr6:38,500,000 to 39,600,000 on hg19 with p12 set to 1 × 10 and 1 × 10□□. Instrument selection rules are in Supplementary Methods.

### 2.6 Grading

Claims were graded across databases with a pre-specified five-level scheme (Confirmed, Consistent, Qualified, Unconfirmed, Opposite). The European arm is descriptive, and the VigiBase arm is graded on directional consistency only.

## 3. Results

### 3.1 The signal is large and it is temporally locked to one date

Against the whole database (12,305,041 reports, 2,007 carrying the NAION term), semaglutide accounted for 879 NAION reports among 79,746 semaglutide reports, ROR 120.8 (95% CI 110.6 to 132.0), IC□□□ 5.23 (Table 1). The other GLP-1 receptor agonists were above background at much smaller magnitude, and SGLT2 inhibitors fell significantly below background at 0.32 (0.12 to 0.84).

Adjusted against metformin, semaglutide reached 76.3 (35.9 to 162.2) with report year in the model and 171.9 (81.1 to 364.1) without it, the year-adjusted figure being the conservative one. The class elevation survives adjustment, with every GLP-1 agent above the metformin reference and SGLT2 inhibitors compatible with no association (Table 1).

**Table 1.** Disproportionality for all drug groups in FAERS against the full deduplicated database, with adjusted RORs against metformin.

| Drug group | Reports | NAION reports | ROR (95% CI) | IC<sub>0-100</sub> | Adjusted ROR vs metformin (95% CI) |
| --- | --- | --- | --- | --- | --- |
| Semaglutide | 79,746 | 879 | 120.8 (110.6 to 132.0) | 5.23 | 76.3 (35.9 to 162.2) |
| Tirzepatide | 156,388 | 137 | 5.7 (4.8 to 6.8) | 1.79 | 7.9 (3.6 to 17.5) |
| Liraglutide | 12,858 | 18 | 8.7 (5.4 to 13.8) | 0.92 | 22.3 (9.0 to 55.0) |
| Dulaglutide | 62,590 | 23 | 2.3 (1.5 to 3.4) | 0.07 | 14.2 (5.6 to 35.6) |
| Exenatide | 18,035 | 1 | 0.34 (0.05 to 2.41) | -4.02 | not estimable |
| Lixisenatide | 3,134 | 0 | 0.98 (0.06 to 15.6) | -4.09 | not estimable |
| SGLT2 inhibitors | 77,206 | 4 | 0.32 (0.12 to 0.84) | -3.12 | 0.42 (0.11 to 1.61) |
| DPP-4 inhibitors | 16,673 | 1 | 0.37 (0.05 to 2.61) | -3.95 | not estimable |
| Metformin | 49,787 | 11 | 1.36 (0.75 to 2.45) | -0.88 | 1 (reference) |
| Atorvastatin | 40,028 | 0 | 0.08 (0.005 to 1.22) | -5.90 | not estimable |

| Drug group | Reports | NAION reports | ROR (95% CI) | IC $\square\square\square$ | Adjusted ROR vs metformin (95% CI) |
| --- | --- | --- | --- | --- | --- |
| **Semaglutide** | 79,746 | 879 | 120.8 (110.6 to 132.0) | 5.23 | 76.3 (35.9 to 162.2) |
| **Tirzepatide** | 156,388 | 137 | 5.7 (4.8 to 6.8) | 1.79 | 7.9 (3.6 to 17.5) |
| **Liraglutide** | 12,858 | 18 | 8.7 (5.4 to 13.8) | 0.92 | 22.3 (9.0 to 55.0) |
| **Dulaglutide** | 62,590 | 23 | 2.3 (1.5 to 3.4) | 0.07 | 14.2 (5.6 to 35.6) |
| **Exenatide** | 18,035 | 1 | 0.34 (0.05 to 2.41) | -4.02 | not estimable |
| **Lixisenatide** | 3,134 | 0 | 0.98 (0.06 to 15.6) | -4.09 | not estimable |
| **SGLT2 inhibitors** | 77,206 | 4 | 0.32 (0.12 to 0.84) | -3.12 | 0.42 (0.11 to 1.61) |
| **DPP-4 inhibitors** | 16,673 | 1 | 0.37 (0.05 to 2.61) | -3.95 | not estimable |
| **Metformin** | 49,787 | 11 | 1.36 (0.75 to 2.45) | -0.88 | 1 (reference) |
| **Atorvastatin** | 40,028 | 0 | 0.08 (0.005 to 1.22) | -5.90 | not estimable |

Ten semaglutide reports named NAION before 3 July 2024 and 869 after it. Among 27,986 pre-alert and 50,881 post-alert semaglutide reports, the odds that a report named NAION were 48.6 times higher in the later period, and 46.8 (24.0 to 109.4) after adjustment. The combined half-yearly rate rose from 9.5 per 10,000 in 2024H1 to 294.6 at its peak in 2025H1 and stood at 190.2 in 2026H1 (Figure 2, Supplementary Table S1.).

**Figure 2.**
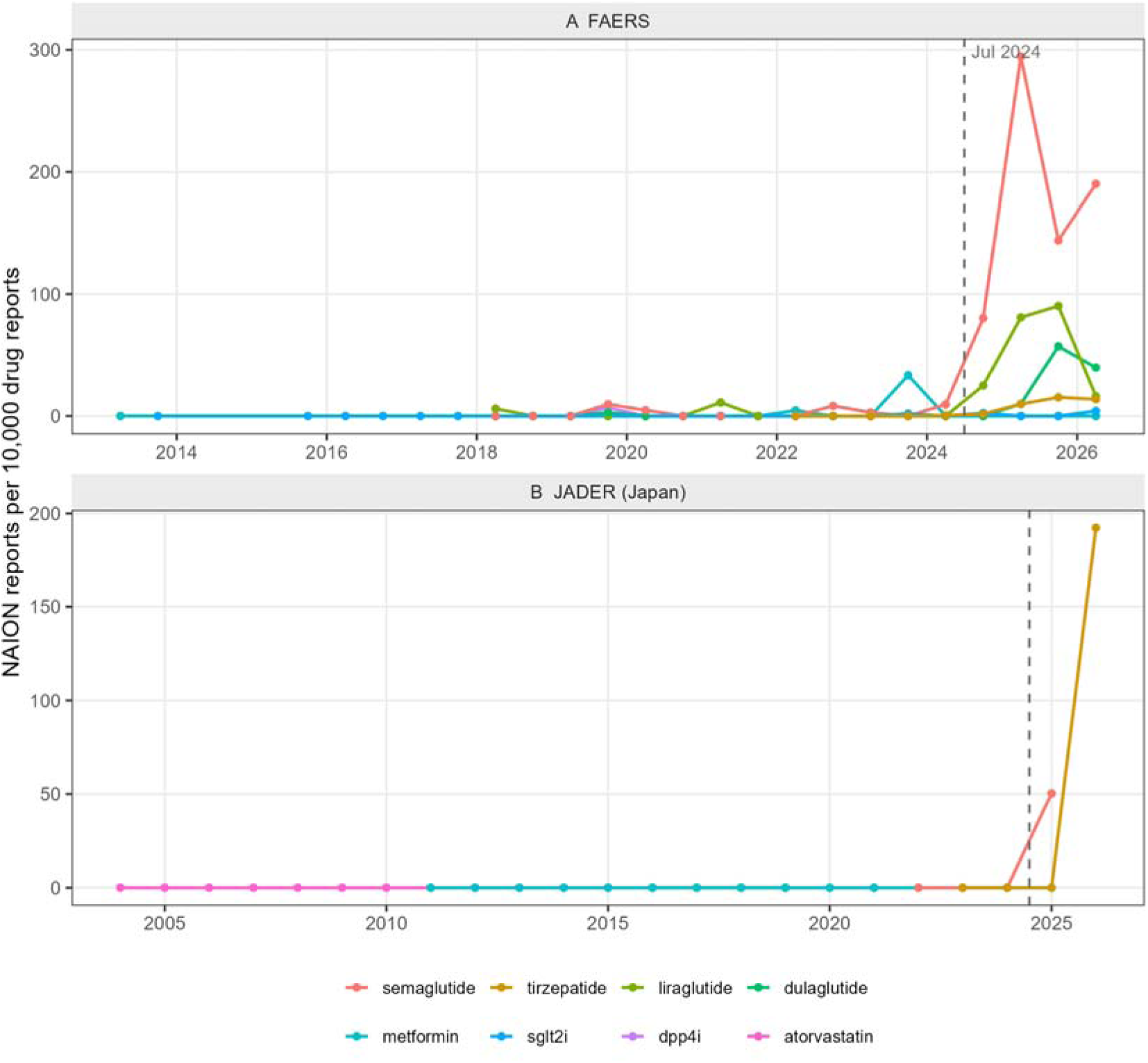
Half-yearly NAION reporting rates per 10,000 drug reports. Panel A shows FAERS from 2018 to 2026 by drug group, with a dashed line at July 2024 marking publication of the index cohort study. Panel B shows JADER from 2004 to 2026, with drug-year cells below 100 reports suppressed to avoid small-denominator artefacts.

### 3.2 The surge is not US-shaped

The post-period by US-region interaction on NAION odds was 0.50 (0.13 to 1.82, P = 0.30). US reports made up 49.3% of semaglutide NAION reports (433 of 879), 6 of 10 before the alert and 427 of 869 after it (P = 0.54).

The regional trajectories then part company (Supplementary Table S1.). The US rate rose from 37.8 per 10,000 in 2024H2 to 224.0 in 2025H1, fell to 81.2, and recovered to 122.7, while the non-US rate rose in every half-year without interruption, 216.2, 384.8, 478.4, and 539.3. A single US-centred publication does not obviously produce a non-US curve still climbing eighteen months later, and the consumer and lawyer share of those reports fell (Supplementary Results).

### 3.3 Onset, reporters, case mix, and the indication background

Median time to onset among 327 semaglutide NAION reports with a computable onset date was 210 days, and the Weibull shape was 0.77 (95% CI 0.71 to 0.85 by bootstrap), an early-decreasing profile that excludes a constant hazard (Figure 3).

**Figure 3.**
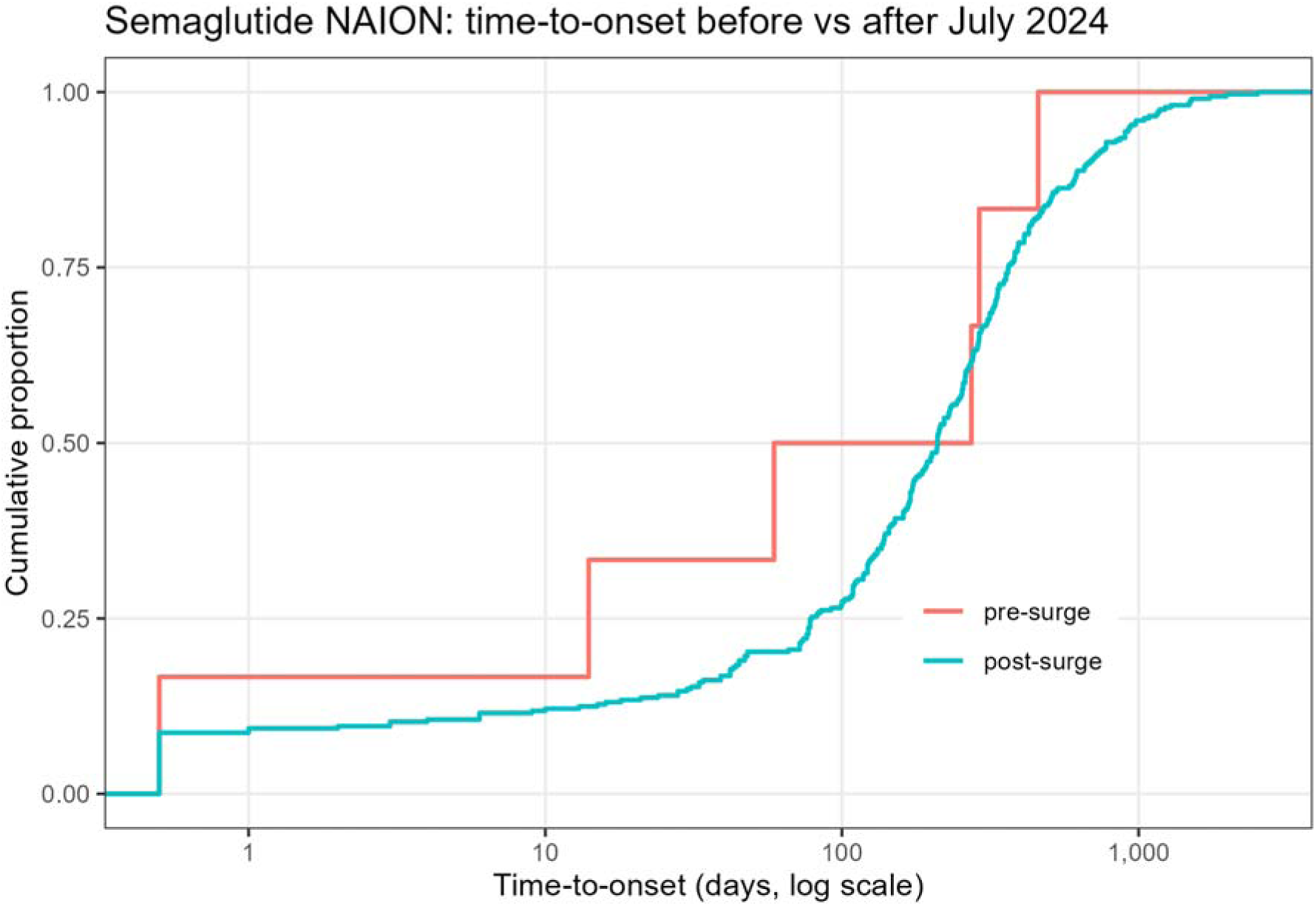
Empirical cumulative distribution of time to onset for semaglutide NAION reports before and after July 2024, on log-scaled days, restricted to reports with a computable onset date. The post-surge distribution is directionally right-shifted (Wilcoxon P = 0.45).

The consumer and lawyer share of all semaglutide reports fell from 0.642 to 0.456 (chi-square P below 0.001), and the same shift appeared in other GLP-1 and in non-GLP-1 antidiabetic reports, so it is system-wide rather than drug-specific. Sex, age-group, and indication-adjusted odds are in Supplementary Results.

Against a background with all GLP-1 reports removed, metformin itself reached an ROR of 5.9 (2.1 to 17.0), so a substantial part of any apparent class effect in this indication area is the diabetes itself. Relative rankings across the two backgrounds were preserved (Spearman ρ = 0.988 for log RORs), and the SGLT2 inhibitor result is the mirror image of a dominant alert depressing its peers.

### 3.4 Analysis of databases of Japan and Europe

JADER contains 1,054,239 reports, 62 carrying the corresponding Japanese term. Semaglutide accounts for two of them among 753 suspect-drug reports and tirzepatide for two among 995. Nothing is gradable at two cases per molecule, but the timing is informative: in a low-awareness environment with no local index study, the first Japanese cases followed the European regulatory action rather than the July 2024 publication. Denmark recorded 28 national reports by 31 May 2025 and 32 by 2 October 2025 [24].

### 3.5 Analysis of VigiBase database

VigiAccess listed 155,748 semaglutide reports on 22 October 2026, of which 871 carried the ischaemic optic neuropathy term, 55.9 per 10,000 reports. Against the study-drug universe the ROR was 24.2 (20.9 to 28.2, q below 0.001), while every other GLP-1 agent fell below expectation (Supplementary Table S2.).

VigiBase holds 871 semaglutide NAION reports where FAERS holds 879, and since US and Japanese reports enter VigiBase through national-centre participation in the WHO Programme, this near-identity means the “global” database is for this association largely the FAERS and JADER reports mirrored. Treating FAERS and VigiBase as two confirmations counts the same reports twice (Supplementary Results).

Within FAERS the elevation is class-wide, while within VigiBase against the study-drug universe liraglutide, dulaglutide, and tirzepatide all sit below 1. FAERS is compared with 12.3 million reports of every kind, VigiBase with a universe in which semaglutide’s own count sets the expectation.

Geographic bounding rules out a regional explanation for the VigiBase pattern. The Americas hold 53.2% of semaglutide reports, so an Americas density advantage requires an Americas share of NAION reports above that figure, and at an assumed 50% share there is none (Supplementary Table S2.).

### 3.6 The genetic instrument is valid, and the ocular results are null

Six variants entered the analysis (Table 2). The maximum pairwise r^2^ is 0.0058, so they are statistically independent despite the shared locus, and the sum of per-variant R^2^ is 0.0083.

The positive control worked: genetically predicted whole-blood GLP1R expression was associated with HbA1c at −0.074 SD per SD of expression (P = 3.0 × 10□□), and the negative control was null (Table 2).

No ocular outcome was associated with the instrument. AION gave an odds ratio of 0.56 per SD of genetically predicted expression (95% CI 0.21 to 1.49, P = 0.25) and glaucoma 1.05 (0.87 to 1.25, P = 0.62), and the four continuous ocular traits gave P of 0.13 or larger (Table 2). Assumption checks were clean (Table 2, Figure 4).

At the observed standard error of 0.496 the AION analysis had 80% power only for an odds ratio above 4.0, and detecting the twofold effect the regulator accepted would require about 980 cases, four times the 244 available. Glaucoma gives 80% power above an odds ratio of 1.29, and the observed 1.05 excludes a doubling.

**Figure 4.**
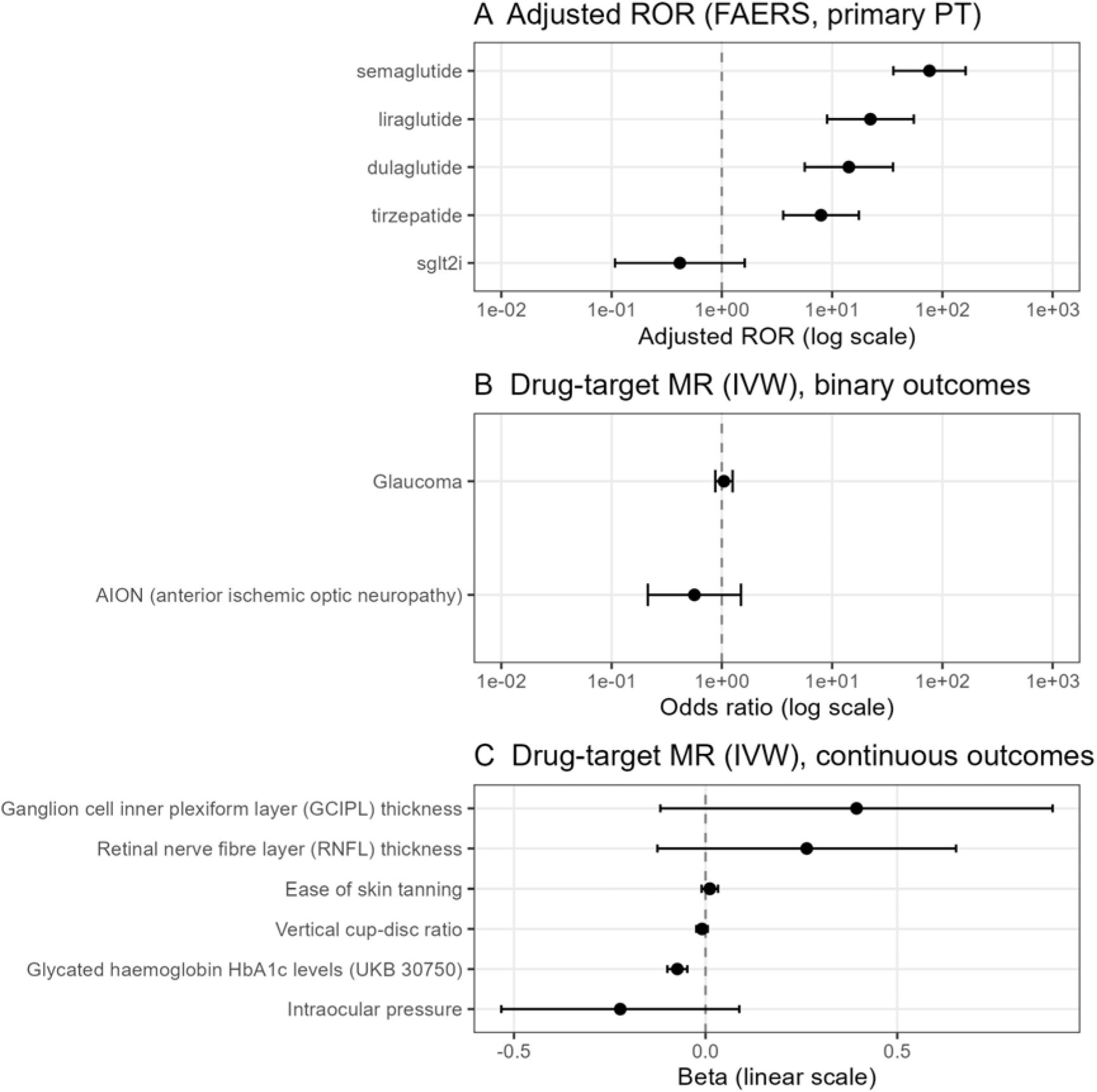
Effect estimates in three panels. Panel A shows covariate-adjusted reporting odds ratios against metformin in FAERS for the primary preferred term, year-adjusted, for drug groups with at least three cases, on a log axis. Panel B shows inverse-variance-weighted drug-target MR odds ratios for the binary outcomes AION and glaucoma, on a log axis with a reference line at 1. Panel C shows the same estimates as per-SD betas for the continuous outcomes, including the HbA1c positive control and the skin tanning negative control, on a linear axis with a reference line at 0.

### 3.7 The colocalisation study

Colocalisation found no shared causal variant between GLP1R expression and any ocular outcome, with PP.H4 at most 0.047 at p12 = 1 × 10 while the expression signal itself was clearly present (PP.H1 between 0.63 and 0.87). The ocular nulls therefore reflect an absence of association with the expression signal rather than instrument failure. For HbA1c and type 2 diabetes, PP.H3 exceeded 0.99, so the metabolic associations in this window are driven by variants distinct from those driving whole-blood GLP1R expression.

A drug-target MR against circulating GLP1R protein would sidestep the tissue question, and it is not available: in the UK Biobank Pharma Proteomics Project Olink Explore data (N = 32,867), 12,921 cis variants produced a minimum P of 4.1 × 10□□.

### 3.8 Grading analysis

The semaglutide excess is Confirmed in FAERS and in VigiBase, with the qualification that VigiBase is not independent, and the comparator-class claim is Qualified because SGLT2 inhibitors fall significantly below background.

Supplementary Table S3. lists every observation that could in principle have distinguished a rare toxicity amplified by awareness from reporting dynamics without toxicity. Of fifteen observations, one favours reporting dynamics, two favour toxicity weakly, two argue against an expression-mediated mechanism, and ten are compatible with both accounts. A literature that reads a single reporting odds ratio as a verdict is reading a quantity that sits among the ten. The rows favouring toxicity are the geography of the step and the falling share of non-US consumer and lawyer reports while the absolute count rose.

## 4. Discussion

Against the three bodies of work summarised in the introduction, this study contributes four things previous work has not. It states the gap between reporting and risk as a quantity, since multi-database studies report both kinds of number and never divide one by the other. It tests the independence the second body assumes rather than granting it, and finds that the agreement between FAERS and VigiBase is the same reports counted twice. It measures how fast a signal can be manufactured, putting a floor under how much of a post-alert estimate is attributable to the alert itself. And it documents instrument independence with a full pairwise LD matrix and adds colocalisation and a protein-level check previous work did not attempt. The contribution is the direction of the test rather than the pairing. The genetic arm sits where it could contradict the reporting arm, a design that transfers to any pair with a reporting signal and a usable cis instrument.

The conclusion belongs to neither camp. The signal is neither established as real nor established as artefact. The reporting quantities in this literature are measurable to three significant figures while the causal quantity is not measurable from these sources at all, for a structural reason rather than a statistical one: spontaneous reporting has no incidence denominator, and the systems that appear to corroborate one another hold the same records.

The MR result is a null with a stated width, and the width matters more than the estimate. The AION odds ratio of 0.56 has an upper limit of 1.49, so the data are compatible with a doubling and exclude only effects above roughly fourfold. The ocular endpoint carries the real information, and if GLP1R expression raised intraocular pressure or produced a glaucoma-like optic neuropathy this design would have seen it. Nothing here excludes a drug action not mediated by whole-blood GLP1R expression at the exposures semaglutide achieves. The colocalisation result is stronger because it does not depend on power in the same way. No ocular trait shares a causal variant with the GLP1R expression signal, and with PP.H3 above 0.99 for HbA1c and type 2 diabetes the metabolic associations in this window are not expression-mediated, so the instrument proxies a quantity of uncertain relevance to the optic nerve head.

The design is pre-specified and hypothesis-neutral, with three reporting systems on different regulatory timelines, two disproportionality backgrounds, masking sensitivity, covariate adjustment, an explicit test of database non-independence, and a genetic layer with colocalisation and a positive control.

VigiAccess gives aggregate counts only, with no deduplication control, no covariates, and no database-wide term totals. There is no NAION-specific preferred term, so the outcome includes other ischaemic optic neuropathies, and 2026 data are partial. JADER contributes two cases per molecule. The instrument set is single-locus, whole-blood, and explains at most 0.83% of expression variance, and the FinnGen AION endpoint has 244 cases and proxies for NAION rather than measuring it. Instruments and outcomes are of European ancestry, and no atlas offers an AION endpoint of comparable size [22, 25]. Residual confounding by indication, diabetes severity, and comorbidity remains in all adjusted models.

Clinicians should continue to act on the labelled risk, and sudden visual loss in a patient on semaglutide needs urgent ophthalmological assessment with withdrawal of the drug if NAION is confirmed [3]. For anyone reading disproportionality outputs, compare the reporting odds ratio with the epidemiological estimate and report the gap. A 121-fold reporting signal against a twofold epidemiological estimate is a 60-fold amplification, and that amplification is a property of the reporting system that will recur with the next high-visibility drug.

## 5. Conclusions

Three numbers carry this paper. The FAERS reporting odds ratio of 120.8 against the relative risk near 2 the regulator accepted is a sixty-fold amplification. The step change of 48.6 within one drug-event pair after a single publication is how fast that amplification is produced. The near-identity of 879 FAERS and 871 VigiBase reports is why the amplification cannot be cross-checked against a second system. Fifteen observations that could have separated toxicity from reporting dynamics separated almost nothing, and the genetic layer bounds one mechanism. The causal question stays open in both directions and will be settled by new-user cohort or self-controlled designs with objective ocular endpoints, not by further disproportionality.

## Supporting information

Supplementary Table S1-S3, Methods and Results

## Declarations

### Data availability

FAERS and JADER are publicly available from the FDA and the PMDA. VigiBase aggregate counts are available through VigiAccess (https://www.vigiaccess.org). Genetic data used for MR and colocalisation are available through OpenGWAS (https://gwas.mrcieu.ac.uk) and eQTLGen (https://www.eqtlgen.org). The code used in this work is available via GitHub at https://github.com/DrWoodWood/Semaglutide-NAION.

### Author contributions

J.J. and D.H. downloaded and analysed the data; J.J., D.H., X.L. and Q.L. wrote the manuscript. Q.Z. and Z.L. conceived and designed this study. Q.Z., L.D. and Z.L. supervised this work and reviewed the manuscript. All authors interpreted results and approved the manuscript.

