## Supplementary Table S1-S3, Methods and Results for "Semaglutide and non-arteritic anterior ischaemic optic neuropathy: a 121-fold reporting signal and a bounded genetic null"

#### 1. Supplementary Table S1. Half-yearly NAION reporting rates per 10,000 semaglutide reports, combined and split by region, for the post-alert period

| **Half-year** | **Combined (NAION / reports)** | **US rate** | **Non-US rate** |
| --- | --- | --- | --- |
| 2024H1 | 9.5 (5 / 5,285) | 2.6 (1 / 3,830) | 27.5 (4 / 1,455) |
| 2024H2 | 80.2 (64 / 7,978) | 37.8 (23 / 6,082) | 216.2 (41 / 1,896) |
| 2025H1 | 294.6 (136 / 4,616) | 224.0 (58 / 2,589) | 384.8 (78 / 2,027) |
| 2025H2 | 144.0 (240 / 16,666) | 81.2 (114 / 14,032) | 478.4 (126 / 2,634) |
| 2026H1 | 190.2 (429 / 22,556) | 122.7 (232 / 18,903) | 539.3 (197 / 3,653) |

#### Supplementary Table S2. VigiBase (VigiAccess) disproportionality against the study-drug universe, retrieved 22 October 2026, with the regional bounding analysis

| **Drug group** | **Reports** | **NAION reports** | **Rate per 10,000** | **ROR (95% CI)** |
| --- | --- | --- | --- | --- |
| Semaglutide | 155,748 | 871 | 55.9 | 24.2 (20.9 to 28.2) |
| Tirzepatide | 233,637 | 144 | 6.2 | 0.56 (0.47 to 0.66) |
| Liraglutide | 76,245 | 25 | 3.3 | 0.31 (0.21 to 0.46) |
| Dulaglutide | 87,389 | 24 | 2.7 | 0.26 (0.17 to 0.39) |
| Metformin | 215,726 | 4 | 0.2 | 0.015 (0.006 to 0.040) |
| Sitagliptin | 37,823 | 0 | 0 | 0.013 (0.001 to 0.20) |
| Empagliflozin | 60,558 | 2 | 0.3 | 0.031 (0.008 to 0.12) |
| Atorvastatin | 215,932 | 16 | 0.7 | 0.060 (0.037 to 0.098) |

Regional bounding for semaglutide, with the Americas holding 82,896 of 155,748 reports (53.2%): assuming 100% of NAION reports come from the Americas gives 105.1 per 10,000 in the Americas and none elsewhere, 75% gives 78.8 against 29.9 (ratio 2.6), and 50% gives 52.5 against 59.8 (ratio 0.9). Tirzepatide holds 139,962 of 233,637 reports in the Americas (59.9%) and 144 NAION reports, giving 10.3 per 10,000 under the 100% assumption.

#### Supplementary Table S3. Every observation that could in principle distinguish a rare toxicity amplified by awareness from reporting dynamics without toxicity

Each row states what each account predicts, what the data showed, and whether the observation separates the two.

| **Observation** | **Toxicity amplified by awareness predicts** | **Reporting dynamics without toxicity predicts** | **Observed** | **Separates the accounts?** |
| --- | --- | --- | --- | --- |
| Step at 3 July 2024 | gradual accumulation, no step at a publication date | a step at the publication | 10 cases in 6.5 years before, 869 in 2 years after, adjusted OR 48.6 (24.0 to 109.4) | Yes, toward reporting dynamics |
| Geography of the step | parallel US and non-US rise | concentrated where the alert was loudest | interaction OR 0.50 (0.13 to 1.82), P = 0.30 | Weakly, toward toxicity |
| Non-US trajectory after 2024H2 | steady once exposure stabilises | a rise at each new alert | US 224.0 then 81.2, non-US 384.8, 478.4, 539.3 | No, the June 2025 European label change predicts the same rise |
| Reporter composition | no change specific to this pair | a system-wide shift toward professionals | 0.642 to 0.456 for semaglutide, 0.867 to 0.737 and 0.383 to 0.123 for comparators | No |
| Consumer and lawyer share of non-US NAION reports | professional reporters find real events | media coverage recruits consumers | share fell from 26.8% to 19.3% while the count rose from 11 to 38 | Weakly, toward toxicity |
| Case mix by sex and age | matches NAION epidemiology | mirrors who files reports | male 2.94, peak at 45 to 64, odds below reference at 75 or more | No, and the oldest stratum contradicts true NAION epidemiology |
| Time to onset | stable onset distribution | case-finding lengthens apparent latency | 210 days after against 166 before, P = 0.45, six pre-surge cases | No |
| Weibull shape of onset | decreasing hazard if a susceptible period exists | constant or increasing hazard under case-finding | semaglutide 0.77 (0.71 to 0.85), tirzepatide 0.74 (0.61 to 0.98), both below 1 | No, and it removes an argument used in earlier work on this pair |
| Indication mix | diabetes and weight management both plausible | prescribing expansion drives report volume | crude 2.1 against 1.8, adjusted weight-management OR 1.11 (0.91 to 1.36) | No |
| Class pattern inside FAERS | a GLP1R class effect | class-wide attention | liraglutide 22.3, dulaglutide 14.2, tirzepatide 7.9 | No |
| Class pattern inside VigiBase | the same class effect | molecule-specific framing | every comparator below 1, from reports overlapping FAERS | No, and it shows the class claim is a property of the background chosen |
| First JADER cases | follow exposure | follow local awareness | 2025 and 2026, after the European action, with no local index study | No |
| MR on AION | elevated if expression-mediated | null | OR 0.56 (0.21 to 1.49), power only above 4.0 | No, uninformative |
| MR on glaucoma | elevated if the optic nerve mechanism is shared | null | OR 1.05 (0.87 to 1.25), power above 1.29 | Yes, against an expression-mediated optic neuropathy |
| Colocalisation at GLP1R | a shared causal variant for any ocular effect | null | PP.H4 at most 0.047 with PP.H1 0.63 to 0.87 | Yes, against expression mediation |

**Summary of the tally.** One observation favours reporting dynamics, two favour toxicity weakly, two argue against an expression-mediated mechanism, and ten are compatible with both accounts. The distribution, rather than any single row, is the finding.

### 2. Supplementary Methods

#### Reporting-dynamic probes, in full

**Regional structure.** Half-yearly NAION rates per 10,000 drug reports were computed separately for US and non-US reports. A logistic model of NAION status on the interaction between a post-period indicator (on or after 3 July 2024) and US region was fitted among semaglutide reports. The post-period odds ratio quoted in the main text comes from the two-by-two table of period against NAION status within semaglutide reports, and the adjusted version adds indication class, sex, and age group.

**Time to onset.** Distributions were compared before and after the surge among reports with a computable onset date, with medians, interquartile ranges, and a Wilcoxon rank-sum test. Weibull shape parameters were fitted where n was at least 20.

**Reporter composition.** Consumer and lawyer shares were computed for semaglutide NAION reports and for all semaglutide reports before and after the alert, and against the same shift in other GLP-1 reports and in non-GLP-1 antidiabetic reports.

**Case mix and indication.** Odds of a NAION mention among semaglutide reports were modelled by sex and age group, before and after addition of indication class and period, and the crude diabetes-versus-weight-management ratio was compared with the ratio among non-NAION semaglutide reports.

**Masking.** Reporting odds ratios were recomputed against a background from which all GLP-1 receptor agonist reports had been removed, and the two sets of log RORs were compared by Spearman correlation.

**Non-US decomposition.** The post-alert non-US rate trajectory was split by reporter type to test whether the increase was accompanied by a growing consumer and lawyer footprint.

**VigiBase geography.** The interface gives region-level report totals per drug but no term-by-region cross-tabulation. NAION reporting density in the Americas was bounded against all other regions under explicit assumptions that 100%, 75%, or 50% of semaglutide NAION reports came from the Americas, with tirzepatide as an internal geographic control. The subset relation between FAERS and VigiBase was checked on the exposure side by comparing the Americas regional total with the FAERS semaglutide total, and on the case side by applying the FAERS NAION density to that Americas exposure.

#### Weibull fitting routes

The shape parameter is fitted twice and the quoted interval is a 2,000-draw percentile parametric bootstrap with seed 20261024, because the two analytic standard errors disagree enough to change the class label.

| **Fitting route** | **semaglutide shape (n = 327)** | **95% CI** | **standard error** | **class** |
| --- | --- | --- | --- | --- |
| Maximum likelihood on the (shape, scale) parameterisation, Hessian standard error | 0.77 | 0.71 to 0.84 | 0.0353 | early-decreasing |
| Accelerated-failure-time reparameterisation with delta-method transform | 0.77 | 0.66 to 0.89 | 0.0576 | early-decreasing |
| Parametric bootstrap, 2,000 draws, 2.5 and 97.5 percentiles | 0.77 | 0.71 to 0.85 | 0.0340 | early-decreasing |

The point estimate and the scale (258.8 days) are identical across all three routes. The accelerated-failure-time standard error is 1.7 times wider because the delta-method transform is poor when shape and scale are strongly correlated. The choice matters: for tirzepatide (n = 45, shape 0.74) the first route gives 0.56 to 0.92 and the second 0.44 to 1.05, which would move the label from early-decreasing to random-type. The bootstrap gives 0.61 to 0.98 and keeps the label.

#### Instrument selection rules

Candidates from chr6:37,600,000 to 39,600,000 (hg19) at P below 1 × 10⁻⁵ in eqtl-a-ENSG00000112164 were retained when the F statistic reached 10 and the per-variant sample size reached 10,000. Palindromic variants were dropped unless the effect allele frequency was reported and the minor allele frequency fell below 0.42. No retained variant was palindromic. F statistics ranged from 19.5 to 77.2 and per-variant sample sizes from 24,479 to 29,621.

### 3. Supplementary Results

#### Time to onset

Among 327 semaglutide NAION reports with a computable onset date, median time to onset was 210 days (interquartile range 78 to 363). Pre-surge reports had a median of 166 days (n = 6) against 210 days post-surge (n = 321), a shift in the case-finding-consistent direction that did not reach significance (Wilcoxon rank-sum P = 0.45, Figure 2). Tirzepatide gave a Weibull shape of 0.74 (95% CI 0.61 to 0.98), also below 1, and onset distributions differed across the GLP-1 agents with at least 20 usable values (Kruskal-Wallis P = 0.0096).

#### Masking against a GLP-1-free background

Against a background from which all GLP-1 receptor agonist reports had been removed, metformin itself reached an ROR of 5.9 (2.1 to 17.0) and semaglutide 127.9 (78.0 to 209.8). Relative rankings across the two backgrounds were otherwise preserved (Spearman ρ = 0.988 for log RORs).

#### Reporter composition and case mix

The consumer and lawyer share of semaglutide NAION reports was 0.300 before the alert and 0.274 after it, which cannot be tested at ten pre-alert reports. Across all semaglutide reports the same share fell from 0.642 to 0.456 (chi-square P below 0.001), against 0.867 to 0.737 in other GLP-1 reports and 0.383 to 0.123 in non-GLP-1 antidiabetic reports.

NAION reports were enriched for diabetes over weight management, a ratio of 2.1 against 1.8 among non-NAION semaglutide reports. Adjustment removed the difference: weight-management indication carried an odds ratio of 1.11 (0.91 to 1.36, P = 0.29) while the post-period effect stayed at 46.8 (24.0 to 109.4), so the expansion of weight-management use does not explain the surge.

Odds of a NAION mention among semaglutide reports were 2.57 (2.15 to 3.07) higher in male than female reports, rising to 2.94 (2.46 to 3.53) once indication and period were added. These are report-level odds, not patient-level risk, and a male predominance fits both the modest male excess reported in incident-NAION series [11] and sex differences in who files reports. Odds peaked at 45 to 64 years (2.38, 1.80 to 3.20, and 2.59, 1.96 to 3.50, fully adjusted), were intermediate at 65 to 74 years (1.34, 0.98 to 1.86, and 1.48, 1.07 to 2.06), and fell below the 18 to 44 reference at 75 years or more (0.67, 0.42 to 1.05, and 0.69, 0.43 to 1.09). The peak sits in the decade of typical NAION onset, while the fall in the oldest stratum runs against the strongly age-graded incidence of NAION and suggests the oldest patients are under-reported rather than protected.

The share of non-US semaglutide NAION reports filed by consumers and lawyers was 26.8% (11 of 41) in 2024H2, then 19.2% (15 of 78), 16.7% (21 of 126), and 19.3% (38 of 197).

#### Japan and Europe in detail

JADER contains 1,054,239 reports, 62 of which carry the corresponding Japanese term. Semaglutide accounts for two of them among 753 suspect-drug reports and tirzepatide for two among 995, with per-report rates close at 2.7 and 2.0 per 1,000. Semaglutide had no NAION report in JADER during 2020 to 2024, the first appearing in 2025 with the second in 2026, and tirzepatide's two both came in 2026. At two cases per molecule nothing is gradable, and the RORs of 46.8 (11.4 to 191.6) for semaglutide and 35.4 (8.6 to 144.8) for tirzepatide are quoted only to show that neither is separable from the other. Estimates for drugs with zero cases are not reported, since the Haldane correction turns them into artefacts of the denominator.

PRAC concluded on 6 June 2025 that NAION is a very rare side effect of semaglutide, up to one in 10,000 patients, drawing on non-clinical, trial, post-marketing, and literature data. Large epidemiological studies indicated an approximately twofold increase in type 2 diabetes, about one additional case per 10,000 person-years [1]. The Danish Medicines Agency recorded 28 national NAION reports with semaglutide by 31 May 2025 (15 Wegovy, 13 Ozempic) and 32 by 2 October 2025 [2]. Product information was updated in September 2025. This arm is descriptive and is not graded.

#### Why the VigiBase count is smaller than the FAERS count

FAERS is a contributor to VigiBase rather than an independent comparator, since the FDA files United States reports with the WHO Programme as a national centre and the PMDA does the same for Japan. For this drug-event pair FAERS is itself only 49.3% United States, with 433 of the 879 NAION reports filed in the US and 446 filed elsewhere.

The exposure side shows the subset relation directly. VigiBase holds 155,748 semaglutide reports, of which 82,896 sit in the Americas, which is 3,150 more than the 79,746 semaglutide reports in FAERS. Applying the FAERS NAION density of 110.2 per 10,000 reports to that Americas exposure predicts 914 NAION reports from the Americas alone, while VigiBase holds 871 in total, so the 72,852 semaglutide reports from every other region contribute almost nothing to the term. An independent global system carrying the same exposure would have been expected to hold about 1,717 NAION reports, and the observed 871 sits 49% below that figure. VigiBase is not running behind FAERS in time, since its 2026 semaglutide count of 48,070 stands at 2.13 times the 22,556 FAERS reports filed in the first half of 2026.

The residual difference of 8 reports, 0.9% of the FAERS count, is the size of gap expected between two extracts of the same material. The FAERS arm is a quarterly ASCII extract covering 2018Q1 to 2026Q2 while VigiAccess is a snapshot taken on 22 October 2026, the two interfaces label the preferred term differently, MedDRA versions differ, FAERS counts report records matched by drug name in the primary-suspect role while VigiAccess counts cases coded to a substance group, and VigiBase merges a case that reaches the Monitoring Centre through more than one centre. VigiAccess publishes aggregates only, so no case-level linkage can settle that difference, and none is required for the claim made here, which is that the global database adds no material information beyond the United States and Japanese records for this association.

#### Colocalisation detail

For AION the posteriors at p12 = 1 × 10⁻⁵ were PP.H1 0.63, PP.H3 0.32, and PP.H4 0.047, which coloc classifies as inconclusive. Across the ocular outcomes PP.H4 was at most 0.047 at p12 = 1 × 10⁻⁵ and at most 0.005 at 1 × 10⁻⁶.

#### Continuous-endpoint power

At 80% power and the observed instrument strength the detectable effects were 0.44 SD on intraocular pressure, 0.021 units on vertical cup-disc ratio, 0.56 SD on retinal nerve fibre layer thickness, and 0.73 SD on ganglion cell inner plexiform layer thickness. For the binary endpoints the AION analysis detected odds ratios above 4.0 and would require about 980 cases to detect an odds ratio of 2, and the glaucoma analysis detected odds ratios above 1.29.

#### Sensitivity estimates for the primary endpoint

For AION the weighted-median estimate was 0.47 (0.14 to 1.51) and the MR-Egger estimate 0.54 (0.02 to 19.2). Leave-one-out refits moved the AION estimate between −0.46 and −0.72 with no P below 0.20. Allowing LD proxies at r² above 0.8 changed nothing for any outcome, because no additional variant was retrieved. For the HbA1c positive control the weighted-median estimate was −0.055 (P = 4.7 × 10⁻⁴), Cochran's Q gave P = 0.20 and the MR-Egger intercept P = 0.18, consistent with a locus carrying expression-independent signals alongside the eQTL.

### Supplementary note on instrument reproducibility

The instrument set was rebuilt on a second machine using a separate PLINK 2 binary and a separate copy of the LDSC 1000 Genomes EUR Phase 3 chromosome 6 reference fileset. The resulting results were identical to the original at six decimal places, with the same six variants, the same maximum pairwise r² of 0.0058, and the same F-statistic range of 19.5 to 77.2.

1.	European Medicines, A. *PRAC concludes eye condition NAION is a very rare side effect of semaglutide medicines Ozempic, Rybelsus and Wegovy*. European Medicines Agency 2025 2025/06/06

2026/10/24; Available from: <https://www.ema.europa.eu/en/news/prac-concludes-eye-condition-naion-very-rare-side-effect-semaglutide-medicines-ozempic-rybelsus-wegovy>.

2.	Danish Medicines, A. *PRAC assesses that NAION could be a very rare side effect of Ozempic, Rybelsus and Wegovy*. Danish Medicines Agency 2025 2025/10/02

2026-10-24; Available from: <https://laegemiddelstyrelsen.dk/en/news/2025/>.
